# DNA Methylation as a Potential Mediator of the Association Between Prenatal Tobacco and Alcohol Exposure and Child Neurodevelopment in a South African Birth Cohort

**DOI:** 10.1101/2022.05.20.22275378

**Authors:** Sarina Abrishamcar, Junyu Chen, Dakotah Feil, Anna Kilanowski, Nastassja Koen, Aneesa Vanker, Catherine J. Wedderburn, Kirsten A. Donald, Heather J. Zar, Dan J. Stein, Anke Hüls

## Abstract

Prenatal tobacco exposure (PTE) and prenatal alcohol exposure (PAE) have been associated with an increased risk of delayed neurodevelopment in children as well as differential newborn DNA methylation (DNAm). However, the biological mechanisms connecting PTE and PAE, DNAm, and neurodevelopment are largely unknown. Here we aim to determine whether differential DNAm mediates the association between PTE and PAE and neurodevelopment at 6 (N=113) and 24 months (N=187) in children from the South African Drakenstein Child Health Study. PTE and PAE were assessed antenatally using urine cotinine measurements and the ASSIST questionnaire, respectively. Cord blood DNAm was measured using the EPIC and 450K BeadChips. Neurodevelopment (cognitive, language, motor, adaptive behavior, socioemotional) was measured using the Bayley Scales of Infant and Toddler Development, Third Edition. We constructed methylation risk scores (MRS) for PTE and PAE and conducted causal mediation analysis (CMA) with these MRS as mediators. Next, we conducted a high-dimensional mediation analysis to identify individual CpG sites as potential mediators, followed by a CMA to estimate the average causal mediation effects (ACME) and total effect (TE).

PTE and PAE were associated with neurodevelopment at 6 but not at 24 months. PTE MRS reached a prediction accuracy (R^2^) of 0.23 but did not significantly mediate the association between PTE and neurodevelopment. PAE MRS was not predictive of PAE (R^2^=0.006). For PTE, 31 CpG sites and 8 CpG sites were identified as significant mediators (ACME and TE *P*<0.05) for the cognitive and motor domains at 6 months, respectively. For PAE, 16 CpG sites and 1 CpG site were significant mediators for the motor and adaptive behavior domains at 6 months, respectively. Several genes including *MAD1L1, CAMTA1*, and *ALDH1A2* have been implicated in neurodevelopmental delay, suggesting that differential DNAm may partly explain the biological mechanisms underlying the relationship between PTE and PAE and child neurodevelopment.

## Introduction

Pregnancy is a critical time window for neurodevelopment and a time when the fetus is most susceptible to adverse environmental and prenatal exposures^1^. Prenatal tobacco exposure (PTE) and prenatal alcohol exposure (PAE) during pregnancy are associated with adverse fetal and childhood outcomes. Maternal smoking and alcohol consumption during pregnancy have been associated with adverse fetal outcomes such as low birth weight, sudden infant death syndrome (SIDS), and preterm birth^2–5^.

PTE and PAE have also been linked to the etiology of a range of neurodevelopmental disorders in children. For example, PTE has previously been associated with attention deficit hyperactivity disorder (ADHD)^6^. There is evidence that the risk of ADHD increases with higher concentrations of PTE^7^. There is also evidence that maternal smoking during pregnancy is a risk factor for neuropsychiatric disorders such as schizophrenia (SZ) and bipolar disorder (BPD)^8,9^. PAE is the cause of fetal alcohol spectrum disorder (FASD), a condition characterized by severe neurodevelopmental delay, and which has various manifestations including growth deficiencies and both behavioral and cognitive deficits^10,11^.

While PTE and PAE have been implicated in the etiology of several neurodevelopmental disorders, the biological mechanisms have yet to be established. Epigenetic modifications, such as differential DNA methylation (DNAm) are dynamic and highly sensitive to external environmental factors^12^. More importantly, DNAm is potentially reversible, indicating that the methylome could be a therapeutic target for disease treatment and prevention^13^.

There is strong evidence from large-scale epigenome-wide association studies (EWAS) suggesting that PTE is associated with changes in the cord blood methylome^14–16^. The evidence is weaker for an association between PAE and the cord blood methylome^17^, although there has been some evidence of epigenomic changes in other tissues such as placental or buccal tissue^18^. While there is evidence that PTE and PAE influence DNA methylation in cord blood, not much is known about the subsequent adverse outcomes, such as delayed neurodevelopment^19,20^ or about the association between newborn DNAm and child neurodevelopment or related disorders in general^21–23^. Thus, the biological mechanisms connecting PTE and PAE, DNAm and child neurodevelopment are largely unknown.

The majority of EWAS have been conducted in high-income countries (HICs) and/or in children of primarily European ancestry. Despite the rapid proliferation of the field of genomics and translational science in the last decade, cohorts from non-European ancestries and low- and middle-income countries (LMICs) are still largely underrepresented^24^. Consequently, there is a large gap in the literature on these underrepresented populations in which risk factors such as low socioeconomic status, poor maternal health, maternal substance use, and adverse child health outcomes including neurodevelopmental delay are at a much higher prevalence^25^.

Here, we investigated whether differential DNAm mediates the relationship between PTE and PAE and neurodevelopment in children of 6 and 24 months of age using data from a well-characterized South African birth cohort, the Drakenstein Child Health Study (DCHS)^26^. For the causal mediation analysis (CMA), we utilized methylation risk scores (MRS) and high-dimensional mediation analysis (HDMA) to increase statistical power and reduce the dimensionality of the methylation data. To our knowledge, this study is the first mediation analysis to examine this relationship and is also one of the first to utilize HDMA and MRS for epigenetic mediation analysis.

## Materials and Methods

### Study population

The DCHS is a South African, population-based birth cohort of African and mixed ancestry and has been previously described ^26^. Our analysis sample consisted of 266 children from the DCHS with DNAm data from cord blood, genotype data, PTE and PAE measures, and important covariates. Of these,113 infants had measurements available from the Bayley Scales of Infant and Toddler Development, third edition (BSID-III) at 6 months of age, and 184 had measurements available from BSID-III at 24 months of age. Mothers were enrolled during their second trimester and followed through pregnancy at two primary clinics, Newman or Mbekweni. Mother-child pairs were then followed from birth until the child was at least 5 years of age. Ethical approval for human subjects’ research was given by the Human Research Ethics Committee of the Faculty of Health Sciences of the University of Cape Town. Written consent for participation was obtained from the mother on behalf of herself and the infant.

### DNA methylation measurements

As described previously, DNA methylation was measured from cord blood using either the Illumina Infinium HumanMethylation450 BeadChip (450K; n=119) or the Infinium MethylationEPIC BeadChip (EPIC; n=147) arrays^23^. Pre-processing was conducted separately for each array in R 3.5.1, using an identical pre-processing pipeline. The 450K and EPIC arrays contained 426,378 probes and 781,536 probes, respectively. The 450k and EPIC arrays were combined using the *minfi* package, which resulted in 316 samples and 453,093 probes available in both arrays^27^. Background subtraction, color correction, and normalization were performed using the *preprocessFunnorm* function^28^. After sample and probe filtering, 273 samples and 409,033 probes remained for analysis. Of these samples, there were 266 complete cases with genotype data, PTE and PAE measures, and important covariates. Batch effects were removed using the *ComBat* function from the *sva* package^29^. Cell type composition estimates were calculated using the most recent cord blood reference dataset^30^.

### Smoking and alcohol measurements

Prenatal tobacco exposure was objectively measured using urine cotinine levels, which were taken within four weeks of enrollment. Urine cotinine was measured using the IMMULITE® 1000 Nicotine Metabolite Kit (Siemens Medical Solutions Diagnostics®, Glyn Rhonwy, United Kingdom)^31^. Urine cotinine levels were classified as non-smoker or passive smoker (<499 ng/ml) or active smoker (≥ 500 ng/ml). The continuous urine cotinine concentrations were used in this analysis^32^.

Prenatal alcohol exposure was measured at the second antenatal study visit, using a dichotomous, composite score calculated from the Alcohol, Smoking, and Substance Involvement Screening Test (ASSIST), a self-report questionnaire, and from retrospectively collected data on alcohol consumption during pregnancy^33^. The ASSIST questionnaire was developed by the World Health Organization and has been validated for use in international settings^34,35^. Details of the composite score calculation have been described elsewhere^33,36,37^.

### Neurodevelopment measurements

Neurodevelopment was assessed at 6 and 24 months of age using the BSID-III ^38,39^. The BSID-III assessment and its composite scores have been previously validated in South African populations ^40,41^. Trained assessors administered the BSID-III to children via direct observation to generate scores for the cognitive, language, and motor development domains. The same trained assessors administered the BSID-III to mothers to report and generate a score for the adaptive behavior and socioemotional domains^33^. Composite scores generated for each domain were used in this analysis.

### Statistical analysis

First, we estimated the association between PTE and PAE and neurodevelopment at age 6 and 24 months in adjusted linear regression analysis. Mediation analyses with differential DNAm as mediators were conducted using MRS and individual CpG sites as mediators. Significant mediators identified at 6 months of age were validated for neurodevelopment measured at the second time point of 24 months.

#### Confounding Assessment

Confounding was assessed by constructing directed acyclic graphs (DAGs). DAGs were created for each exposure-mediator (E-M) (PTE/PAE-DNAm), mediator-outcome (M-O) (DNAm-neurodevelopment), and exposure-outcome (E-O) relationship (PTE/PAE-neurodevelopment) (Figure S1). Potential confounders were selected based on existing literature^33^. All models in this analysis were adjusted for maternal age, maternal HIV status, maternal depression, maternal psychological distress, parental socioeconomic status (SES), gestational age, and cell-type proportions. Population stratification was adjusted for using the first five genetic principal components. Models evaluating PTE as the primary exposure were additionally adjusted for PAE and likewise, models evaluating PAE as the primary exposure were additionally adjusted for PTE.

#### Methylation Risk Scores

First, we constructed methylation risk scores (MRS) for PTE and PAE as described elsewhere^42^. All 266 samples with DNAm data were included in the MRS calculation. MRS are calculated as the weighted sum of methylation beta levels of CpG sites. MRS calculations were based on summary statistics from epigenome-wide association studies (EWAS) for DNAm probes in which the association between the exposure and DNAm was investigated. These summary statistics, which describe the effect size (beta) at each CpG site in the respective EWAS, were used as external weights to calculate the methylation risk score in our cohort. External summary statistics were acquired from EWASs conducted by the Pregnancy and Childhood Epigenetics (PACE) consortium. PTE summary statistics were acquired from an EWAS investigating the association between maternal smoking and cord-blood DNAm (N=5648) ^16^. PAE summary statistics were acquired from an EWAS investigating the association between maternal alcohol use and cord-blood DNAm (N=1147) ^17^. Both EWASs were comprised of cohorts from primarily European ancestry. To correct for correlated CpG sites, we estimated co-methylated regions (CMR) using the *CoMeBack* package ^43^. Then, we conducted “clumping”, in which one CpG site with the smallest p-value from the external summary statistics per CMR was included in the final MRS. MRS was calculated at several p-value thresholds for each EWAS, a procedure similar to “thresholding” in traditional polygenic risk score calculations. Finally, we calculated the correlation between the exposure and the resulting MRS at different p-value thresholds and chose the p-value threshold associated with the highest prediction accuracy for the subsequent analyses.

#### High-Dimensional Mediation Analyses (HDMA)

MRS are limited in that they only consider the E-M but not the M-O relationship. HDMA considers the E-M as well as the M-O relationship, which allows for a more holistic analysis of the mediating relationships. We performed two separate high-dimensional mediation analyses using the R packages *HIMA* and *DACT* for any exposure-outcome combinations for which we identified indications of a total effect ^44,45^. The *HIMA* package utilizes a penalized-based regression and consists of three steps. First, it performs dimension reduction through sure independence screening to identify the n/log(n) CpG sites with the largest effect size in the mediator-outcome regression model. Second, the minimax concave penalty is applied to this subset of CpG sites for further dimension reduction^46^. Finally, a joint significance test is conducted to evaluate the significance of the mediation effects using a Benjamini-Hochberg false discovery rate (FDR) correction for multiple testing. The divide-aggregate composite null test (DACT) is a more recent method for high-dimensional mediation analysis that is better powered than HIMA ^45^. First, it utilizes the Efron empirical null framework to estimate the proportions of the three null cases across all epigenome mediators ^47^. Then, it performs the DACT for the composite null of no mediation effect in three cases in which the M-O effect is non-zero, the E-M effect is non-zero, or both effects are non-zero. Finally, the DACT p-value is calculated as the weighted sum of all three p-values under the three-null hypothesis. We conducted an association analysis for the E-M and M-O associations with robust linear regression using the *rlm* function to input into DACT^48^. We then pre-filtered CpG sites by two criteria to alleviate the burden of multiple testing. First, we filtered the CpG sites by a p-value threshold of 0.05 for the E-M and M-O models. Second, we filtered CpG sites by the direction of the E-M and M-O effects to achieve an overall negative indirect effect, in line with the hypothesized negative total and direct effects. After running DACT for the resulting subset of CpGs, we corrected for multiple testing using the Benjamini-Hochberg FDR method.

#### Causal Mediation Analysis (CMA)

Finally, we conducted a CMA for the MRS and significant CpG sites from HDMA as mediators. The *mediation* R package was used to estimate mediation effects ^49^. The *mediation* package estimates the average causal mediation effect (ACME), the average direct effect (ADE), the total effect (TE), and the proportion mediated (PM) in the population. 95% confidence intervals were constructed using a nonparametric, bootstrapped, quasi-Bayesian method ^50^. To examine trends across both time points, we cross-validated significant CpG sites identified at 6 months by determining whether they are also significant mediators of the association at 24 months of age.

## Results

### Study population characteristics

This analysis sample included 266 children with DNAm data, genotype data, and other relevant covariates, with a subset of 113 children and 184 children with information on neurodevelopment across 5 domains, respectively (Table 1). This is a cohort of children from African (55%) and mixed (45%) ancestry. There was a high prevalence of maternal smoking with 47% of mothers classified as passive smokers and 30% classified as active smokers. The prevalence of prenatal alcohol use among mothers was 17%. Additionally, 25% of mothers were classified as being above the Beck Depression Inventory II threshold and 30% being at elevated risk of psychological distress on the Self-Reporting Questionnaire (SRQ-20). The prevalence of mothers with an HIV diagnosis was 24%, but all children in this population remained uninfected. Among the subset of children with information on neurodevelopment at 6 months of age, the prevalence of mothers who smoked and reported alcohol use was higher compared to the whole population with DNAm data and children at 24 months, with 35% of mothers classified as active smokers and 23% of mothers classified as consuming alcohol.

**Table 1:** Study population characteristics for the whole sample, children with BSID-III measurements at 6 months of age, and children with BSID-III measurements at 24 months of age

|  | Whole study sample | Children with BSID-III data at 6 mo | Children with BSID-III data at 24 mo |
| --- | --- | --- | --- |
| **N** | 266 | 113 | 184 |
| **Maternal Age (yrs) (median [IQR])** | 26 [22,31] | 25 [22,31] | 26 [22,32] |
| **Gestational Age (wks) (median [IQR])** | 39 [38,40] | 39 [38,40] | 39 [38,40] |
| **Child Sex (%)** |  |  |  |
| Female | 119 (44.74) | 54 (47.79) | 82 (43.85) |
| Male | 147 (55.26) | 59 (52.21) | 105 (56.14) |
| **Parental SES<sup>1</sup> (%)** |  |  |  |
| Lowest SES | 66 (24.81) | 23 (20.35) | 42 (22.46) |
| Low-moderate SES | 65 (24.44) | 33 (29.20) | 47 (25.13) |
| Moderate-high SES | 74 (27.82) | 30 (26.55) | 50 (26.74) |
| Highest SES | 61 (22.93) | 27 (23.89) | 48 (25.67) |
| **Ancestry (%)** |  |  |  |
| Black African | 147 (55.26) | 54 (47.79) | 96 (51.34) |
| Mixed | 119 (44.74) | 59 (52.21) | 91 (48.66) |
| **Maternal Alcohol Use (%)** |  |  |  |
| Non-alcohol user | 219 (82.33%) | 87 (77.00) | 151 (80.75) |
| Alcohol user | 46 (17.29%) | 26 (23.00) | 35 (18.72) |
| **Maternal Smoking<sup>2</sup> (%)** |  |  |  |
| Non-smoker | 60 (22.56) | 24 (21.24) | 44 (23.53) |
| Passive Smoker | 126 (47.37) | 48 (42.48) | 80 (42.78) |
| Active Smoker | 79 (29.70) | 40 (35.40) | 63 (33.69) |
| **Maternal Smoking (ng/ml) (median [IQR])** | 38.40 [11.4, 500.00] | 57.35 [11.48, 500.00] | 55.90 [10.80, 500.00] |
| **Maternal HIV (%)** |  |  |  |
| Yes | 64 (24.06) | 29 (25.66) | 43 (23.00) |
| No | 202 (75.94) | 84 (74.34) | 144 (77.00) |
| **Beck Depression Inventory Threshold (%)** |  |  |  |
| Above | 67 (25.19) | 36 (31.86) | 48 (25.67) |
| Below | 196 (73.68) | 75 (66.37) | 136 (72.73) |
| **Self-Reporting Questionnaire (%)** |  |  |  |
| High Risk | 80 (30.08) | 34 (30.09) | 58 (31.02) |
| Low Risk | 184 (69.17) | 78 (69.03) | 127 (67.91) |
| **Bayley Scales 6 months (median [IQR])** |  |  |  |
| Cognitive (n=113) | - | 105 [95,110] | - |
| Language (n=109) | - | 103 [90.5,112] | - |
| Motor (n=110) | - | 112 [100,118] | - |
| Adaptive Behavior (n=112) | - | 102 [96,107] | - |
| Social-Emotional (n=112) | - | 115 [100,125] | - |
| **Bayley Scales 24 months (median [IQR])** |  |  |  |
| Cognitive (n=184) | - | - | 85 [80,90] |
| Language (n=173) | - | - | 83 [74,91] |
| Motor (n=173) | - | - | 91 [85,100] |
| Adaptive Behavior (n=184) | - | - | 84 [71,92] |
| Social-Emotional (n=184) | - | - | 115 [100,135] |

<sup>1</sup>Socioeconomic status (SES) composite scores (see Supplemental for details) <sup>2</sup>Urine cotinine levels (ng/ml) classified as non-smoker (<10 ng/ml); passive smoker (10-499 ng/ml); active smoker (≥ 500 ng/ml)

### The total effect of prenatal smoking and alcohol exposure on child neurodevelopment

The total effects of PTE and PAE on child neurodevelopment across 5 domains were estimated using adjusted linear regression (Figure 1). There was a consistent negative association between PTE and neurodevelopment across all domains at 6 months of age, which was significant for the cognitive domain (β=-0.02; 95% CI: -0.028, 0.002; *P*= 0.023). There was also a consistent negative association between PAE and neurodevelopment across all domains at 6 months of age, which was significant for the motor domain (β=-9.36; 95% CI: -16.64, -2.08; *P*= 0.012). No associations were found between PTE and PAE and neurodevelopment at 24 months in this sample (Figure S2; Table S1-S2). Therefore, the subsequent mediation analyses focused on neurodevelopment at 6 months as the primary outcome. Neurodevelopment at 24 months was used as validation for any significant mediators identified at age 6 months.

**Figure 1:**
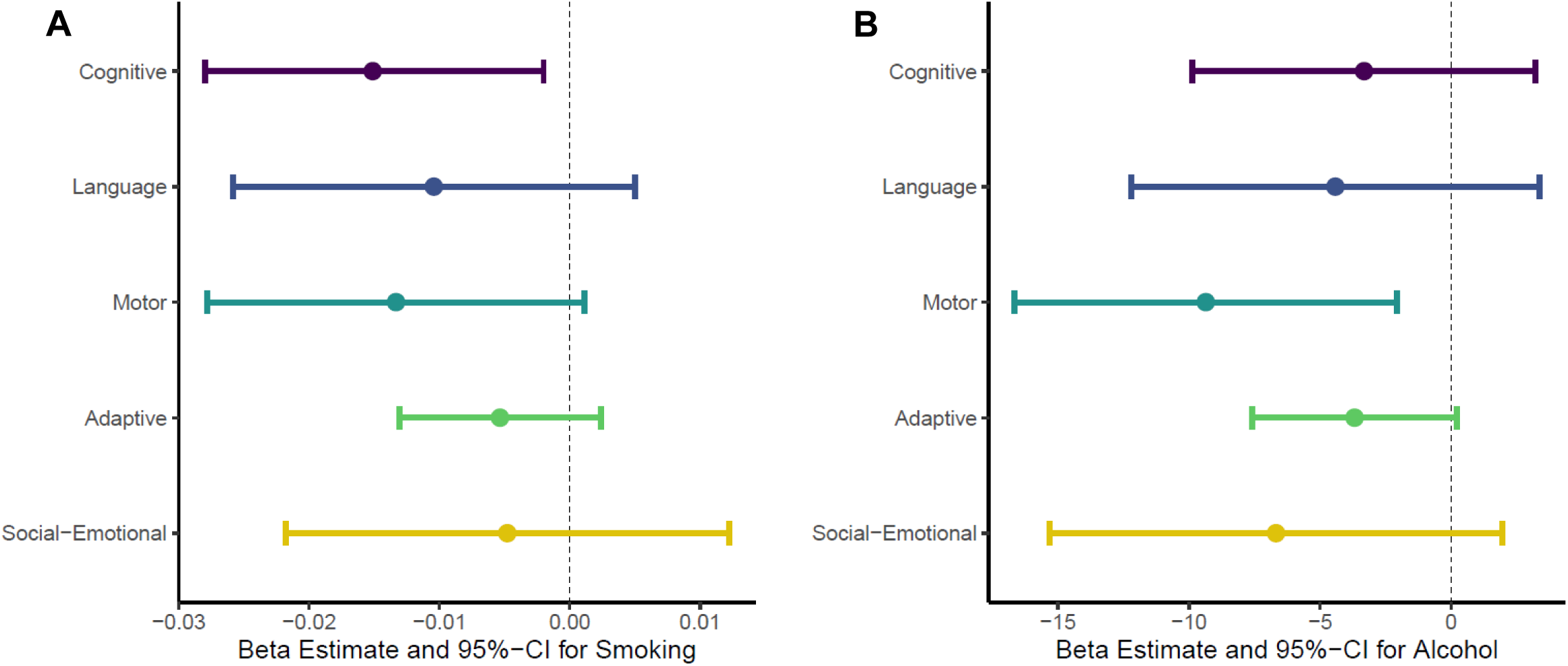
Association of PTE and PAE on neurodevelopment domains at 6 months. Models were adjusted for paternal SES, maternal depression, maternal psychological distress, gestational age, maternal age, maternal HIV status, cell-type proportions, and the first 5 genetic principal components. **1A)** Association of PTE on neurodevelopment domains, additionally adjusted for PAE. **1B)** Association of PAE on neurodevelopment domains, additionally adjusted for PTE.

### Mediation analyses with methylation risk scores as mediators

The MRS for PTE was well correlated with prenatal cotinine levels, with the highest R^2^being 0.23 at a p-value threshold of 5e-21 (Figure 2A). As evident from Figure 2B, active smokers had a higher MRS than non-smokers and passive smokers combined. Additionally, there was a strong and significant association between PTE and the MRS (β=0.0022; 95% CI: 0.0016, 0.0027; *P*=6.7E-15) (Figure 2C). In contrast, the MRS for PAE was not correlated with alcohol use status, with the highest R^2^ being 0.006 at a p-value threshold of 5e-06 (Figure 2D). There was no difference between the MRS for children with and without alcohol exposure (Figure 2E) and there was no association between PAE and the MRS (β=5.4e-03; 95%CI: -0.26,0.27; *P*=0.97) (Figure 2F). Because of the poor performance of the PAE methylation risk score, we did not proceed with CMA for PAE using this approach.

**Figure 2:**
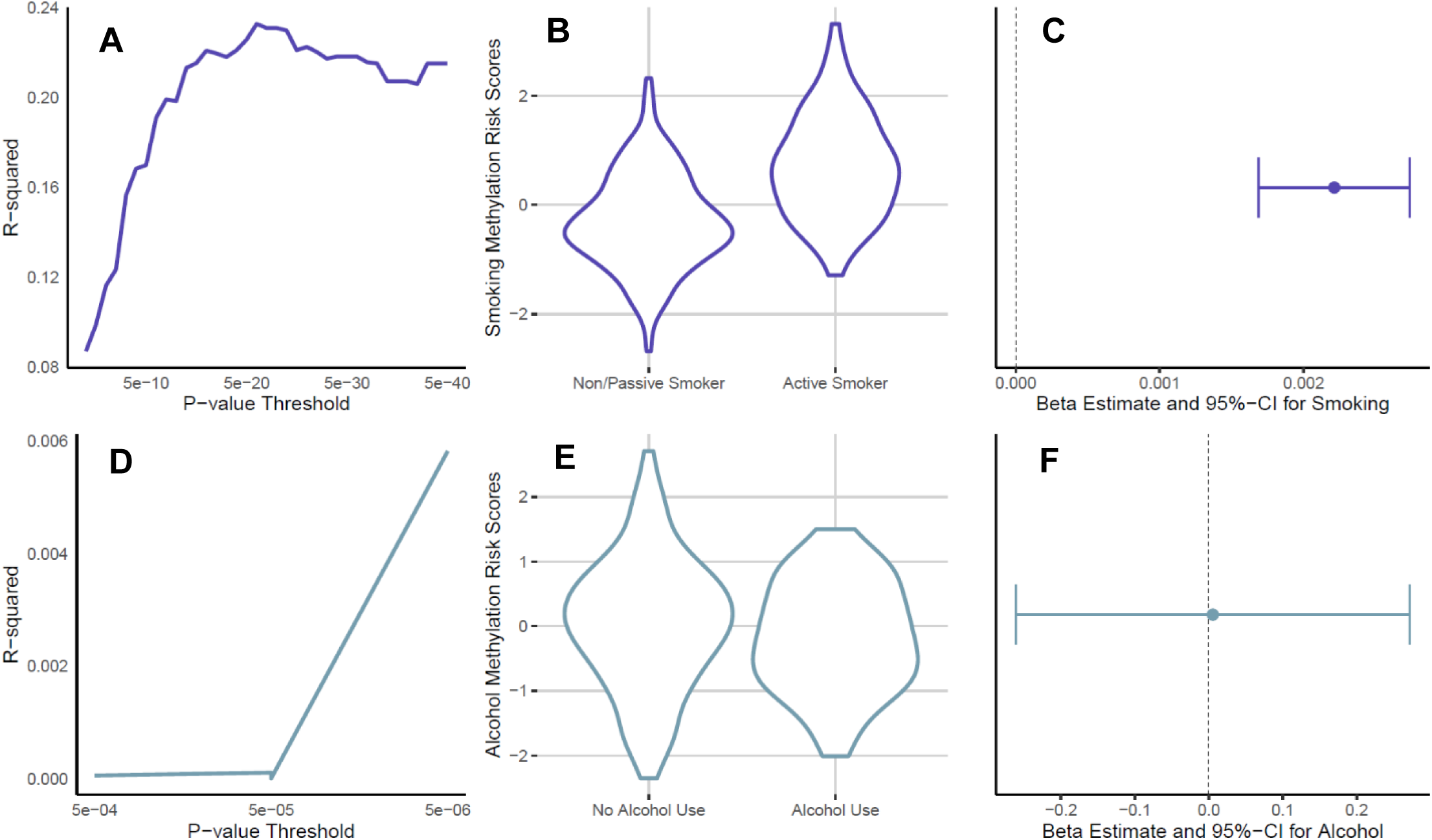
Performance of the MRS for PTE and PAE. 2A) Correlation between PTE and MRS at several p-value thresholds **2B)** MRS for non-/passive smoking mothers and active smoking mothers. **2C)** Effect of PTE on most highly correlated MRS. **2D)** Correlation between PAE and MRS at several p-value thresholds **2E)** MRS for non-alcohol consuming mothers and alcohol consuming mothers **2F)** Effect of PAE on most highly correlated MRS

There was no evidence of mediation by MRS in any domain (Figure 3; Table S3). While the cognitive domain showed a significant total effect, a significant indirect effect (ACME) was not found (ACME=0.006; 95%CI: -3.84E-03, 0.002; *P*=0.2). The direct effect was significant, however (ADE=-0.02; 95%CI: -0.04, -0.01; *P*=0.01).

**Figure 3:**
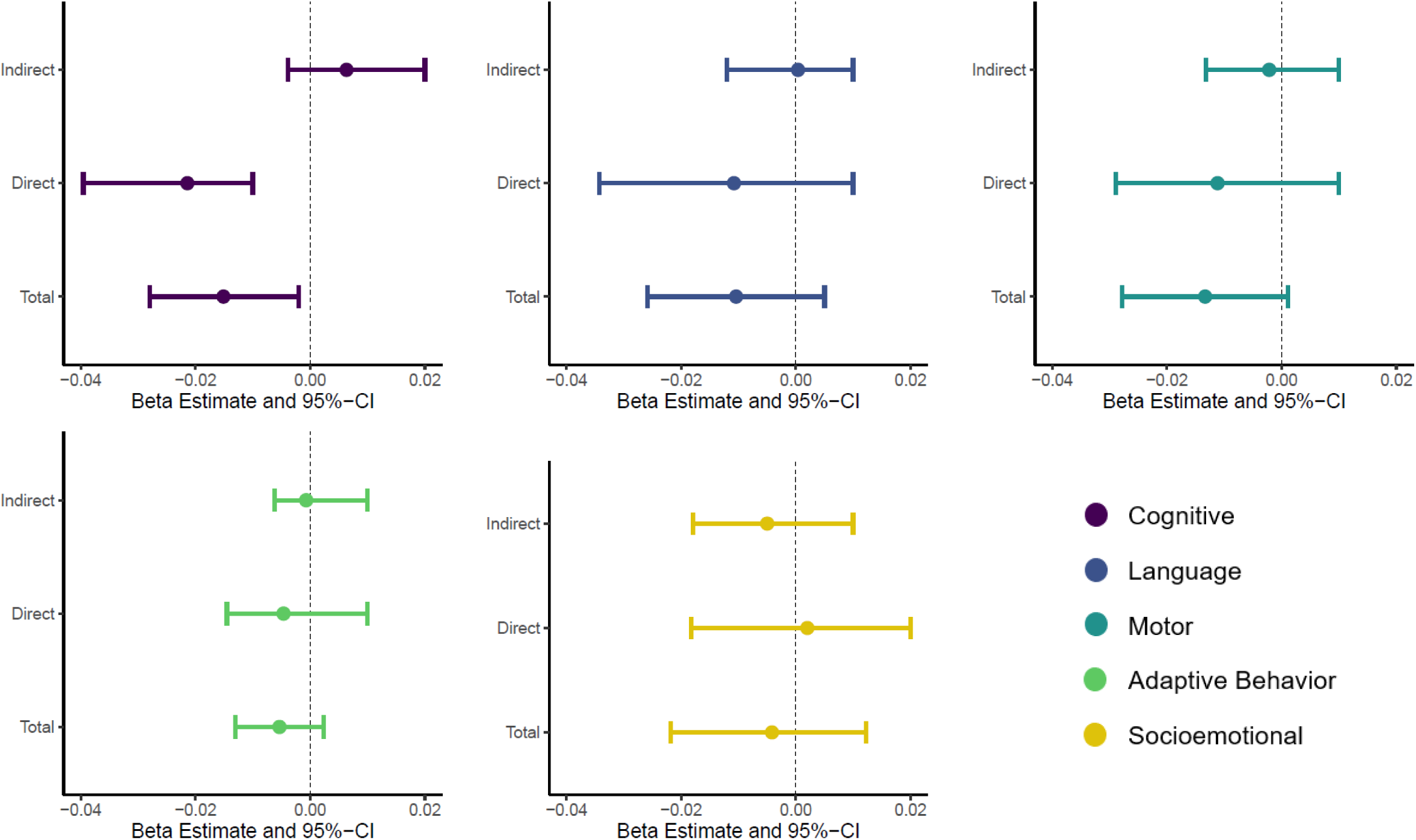
Causal mediation analysis for PTE as the exposure, MRS as the mediator, and neurodevelopment across 5 domains at 6 months of age as outcome. Estimated indirect effect, direct effect, and total effect with their corresponding 95% confidence intervals are shown.

### HDMA with individual CpG sites as mediators

HDMA was conducted for exposure-outcome combinations for which there was an indication of a total effect: PTE-cognitive, PTE-motor, PAE-motor, and PAE-adaptive behavior. After correcting for multiple testing, DACT identified a total of 381 CpG sites as significant mediators of the association between PTE and neurodevelopment across the cognitive (123 CpG sites), language (94 CpG sites), motor (68 CpG sites), adaptive behavior (61 CpG sites), and socioemotional (35 CpG sites). After validating these CpG sites with CMA, we identified 31 CpG sites to have a significant mediating effect (ACME *P* and TE *P* < 0.05) between PTE and cognitive development and 8 CpG sites to have a significant mediating effect between PTE and motor development (Table 2). Three of these CpG sites overlapped between both the cognitive and motor domains (cg22263591 [*SLC39A11*], cg25284031 [*HVCN1*], cg25857569 [*PPM1L*]).

**Table 2:** Significant CpG sites (ACME *P* and TE *P* <0.05) resulting from HDMA and CMA of DNAm as a potential mediator of the association between PTE and neurodevelopmental delay at 6 months of age using the DACT method and after correction for multiple testing (BH FDR ≤0.05). ACME, ADE, TE, and PM effect sizes and 95% confidence intervals estimated by CMA.

| Domain | CpG (Gene) | ACME (95% CI) | ADE (95% CI) | Total Effect (95% CI) | Prop Med. (95% CI) |
| --- | --- | --- | --- | --- | --- |
| Cognitive | cg00480142 | -0.004 (-8.9e-03, -5.0e-05) | -0.011 (-0.025, 2.4e-03) | -0.015 (-0.027, -2.4e-03) | 0.24 (-0.012, 1.4) |
|  | cg00613752 (GPR6) | -0.004 (-9.2e-03, -1.9e-04) | -0.012 (-0.024, 4.2e-04) | -0.016 (-0.029, -3.4e-03) | 0.25 (3.2e-03, 0.88) |
|  | cg01258793 (MAD1L1) | -0.005 (-0.012, -3.4e-04) | -0.011 (-0.024, 2.3e-03) | -0.016 (-0.030, -2.7e-03) | 0.31 (5.5e-03, 1.3) |
|  | cg02215141 | -0.004 (-9.6e-03, -5.5e-05) | -0.011 (-0.024, 2.5e-03) | -0.016 (-0.028, -2.5e-03) | 0.28 (-0.022, 1.3) |
|  | cg02423105 (CDH26) | -0.005 (-0.012, -6.0e-05) | -0.014 (-0.026, -1.2e-03) | -0.018 (-0.031, -4.8e-03) | 0.26 (1.1e-03, 0.83) |
|  | cg02668773 (MPO) | -0.007 (-0.014, -7.7e-04) | -0.012 (-0.025, 1.4e-03) | -0.019 (-0.033, -4.8e-03) | 0.36 (0.05, 1.1) |
|  | cg02879122(CCDC144A) | -0.004 (-9.3e-03, -3.8e-04) | -0.012 (-0.025, 1.2e-03) | -0.016 (-0.028, -2.9e-03) | 0.26 (0.018, 1.2) |
|  | cg03288340 (ZC3H4) | -0.005 (-0.012, -6.3e-04) | -0.011 (-0.023, 2.1e-03) | -0.016 (-0.029, -3.2e-03) | 0.33 (0.039, 1.3) |
|  | cg05129050 (CAMTA1) | -0.005 (-0.011, -6.9e-04) | -0.010 (-0.023, 4.5e-03) | -0.014 (-0.027, -1.4e-03) | 0.35 (9.1e-03, 2.0) |
|  | cg05331340 (ASGR1) | -0.010 (-0.020, -1.8e-03) | -0.012 (-0.025, 7.9e-04) | -0.022 (-0.037, -7.6e-03) | 0.44 (0.093, 1.0) |
|  | cg08481075 (COL11A2) | -0.005 (-0.011, -2.9e-04) | -0.012 (-0.024, 1.9e-03) | -0.016 (-0.029, -2.9e-03) | 0.28 (2.5e-03, 1.1) |
|  | cg08836481 (LARP4B) | -0.008 (-0.016, -1.2e-03) | -0.012 (-0.025, 4.2e-04) | -0.020 (-0.034, -6.2e-03) | 0.39 (0.08, 1.0) |
|  | cg09115275 (IL2RA) | -0.004 (-8.3e-03, -2.2e-04) | -0.012 (-0.024, 1.5e-03) | -0.015 (-0.027, -2.3e-03) | 0.23 (-0.015, 1.1) |
|  | cg09964326 (FGFBP3) | -0.004 (-0.010, -4.5e-04) | -0.012 (-0.025, 1.2e-03) | -0.017 (-0.030, -3.3e-03) | 0.27 (0.024, 1.2) |
|  | cg10149329 (MFSD8) | -0.004 (-9.2e-03, -3.7e-04) | -0.011 (-0.024, 3.0e-03) | -0.015 (-0.027, -1.9e-03) | 0.27 (-2.7e-03, 1.4) |
|  | cg10780097 (AMD1) | -0.003 (-8.0e-03, -1.6e-05) | -0.012 (-0.024, -5.9e-04) | -0.015 (-0.028, -2.6e-03) | 0.20 (-5.9e-03, 0.76) |
|  | cg12031962 (ALDH1A2) | -0.004 (-0.010, -3.5e-04) | -0.011 (-0.024, 3.1e-03) | -0.016 (-0.029, -2.5e-03) | 0.26 (0.015, 1.4) |
|  | cg12428160 | -0.005 (-0.014, -4.3e-04) | -0.013 (-0.025, 7.9e-04) | -0.018 (-0.031, -4.0e-03) | 0.28 (0.025, 1.1) |
|  | cg13889963 (CALB1) | -0.004 (-9.4e-03, -8.2e-05) | -0.011 (-0.024, 2.1e-03) | -0.015 (-0.028, -2.2e-03) | 0.28 (-0.039, 1.2) |
|  | cg13918314 (ANAPC13) | -0.004 (-9.8e-03, -2.1e-05) | -0.013 (-0.025, -6.1e-04) | -0.017 (-0.030, -3.4e-03) | 0.22 (-5.5e-03, 0.84) |
|  | cg14323109 (KDR) | -0.004 (-9.1e-03, -8.4e-05) | -0.011 (-0.023, 3.1e-03) | -0.015 (-0.027, -9.5e-04) | 0.24 (-0.01, 1.6) |
|  | cg14896134 | -0.007 (-0.016, -6.0e-04) | -0.010 (-0.023, 2.0e-03) | -0.017 (-0.030, -3.8e-03) | 0.40 (0.035, 1.2) |
|  | cg15624376 (HOXA4) | -0.004 (-0.010, -2.2e-04) | -0.012 (-0.025, 7.9e-04) | -0.016 (-0.029, -2.6e-03) | 0.25 (1.7e-03, 1.1) |
|  | cg18780259 (GDF1) | -0.005 (-0.012, -5.1e-04) | -0.010 (-0.022, 2.8e-03) | -0.015 (-0.028, -2.3e-03) | 0.34 (0.022, 1.3) |
|  | cg19611886 | -0.009 (-0.021, -8.2e-04) | -0.012 (-0.025, 1.6e-03) | -0.021 (-0.035, -6.2e-03) | 0.42 (0.034, 1.1) |
|  | cg22263591 (SLC39A11) | -0.006 (-0.014, -3.9e-04) | -0.013 (-0.025, -4.9e-04) | -0.019 (-0.031, -5.1e-03) | 0.33 (0.022, 0.89) |
|  | cg23219570 (FGF23) | -0.007 (-0.016, -4.5e-04) | -0.011 (-0.024, 1.5e-03) | -0.019 (-0.032, -5.6e-03) | 0.40 (0.021, 1.1) |
|  | cg23643526 (HAO2) | -0.004 (-9.5e-03, -1.3e-04) | -0.011 (-0.024, 3.0e-03) | -0.015 (-0.027, -2.2e-03) | 0.25 (-0.022, 1.4) |
|  | cg25284031 (HVCN1) | -0.006 (-0.015, -2.2e-04) | -0.014 (-0.026, -5.0e-04) | -0.019 (-0.033, -5.8e-03) | 0.30 (9.9e-03, 0.91) |
|  | cg25823085 (ABCB4) | -0.004 (-9.1e-03, -2.0e-04) | -0.011 (-0.023, 2.2e-03) | -0.014 (-0.027, -1.5e-03) | 0.27 (-0.021, 1.2) |
|  | cg25857569 (PPM1L) | -0.007 (-0.015, -1.6e-04) | -0.012 (-0.025, 5.7e-05) | -0.019 (-0.033, -5.1e-03) | 0.36 (5.5e-03, 0.94) |
| Motor | cg00328376 (SKAP2) | -0.010 (-0.03, -4.9e-03) | -0.010 (-0.023, 9.0e-03) | -0.023 (-0.042, -4.6e-03) | 0.65 (0.26, 2.1) |
|  | cg09490277 (FANCE) | -0.010 (-0.024, -5.3e-04) | -0.011 (-0.026, 4.4e-03) | -0.022 (-0.043, -3.1e-03) | 0.49 (0.019, 1.4) |
|  | cg14989316 (ZMIZ1-AS1) | -0.008 (-0.017, -1.6e-03) | -0.010 (-0.025, 6.5e-03) | -0.018 (-0.035, -1.9e-04) | 0.46 (-7.2e-03, 2.9) |
|  | cg15188623 (ZNF710) | -0.008 (-0.021, -6.8e-04) | -0.010 (-0.025, 6.0e-03) | -0.018 (-0.037, -8.3e-04) | 0.46 (-0.015, 2.1) |
|  | cg22263591 (SLC39A11) | -0.006 (-0.014, -2.8e-05) | -0.011 (-0.026, 4.8e-03) | -0.017 (-0.032, -3.6e-04) | 0.35 (-0.074, 2.0) |
|  | cg24671734 (BTBD11) | -0.007 (-0.019, -6.7e-04) | -0.011 (-0.027, 6.1e-03) | -0.018 (-0.034, -1.9e-03) | 0.41 (-0.014, 2.0) |
|  | cg25284031 (HVCN1) | -0.007 (-0.017, -3.1e-04) | -0.012 (-0.026, 4.6e-03) | -0.018 (-0.035, -1.9e-03) | 0.36 (-0.037, 1.8) |
|  | cg25857569 (PPM1L) | -0.010 (-0.023, -2.9e-03) | -0.010 (-0.024, 6.8e-03) | -0.020 (-0.037, -2.6e-03) | 0.57 (0.17, 1.9) |

DACT identified 395 CpG sites as significant mediators of the association between PAE and neurodevelopment across the cognitive (79 CpG sites), language (74 CpG sites), motor (100 CpG sites), adaptive behavior (93 CpG sites), and socioemotional (49 CpG sites) domains. After conducting CMA, this resulted in 16 CpG sites and 1 CpG site with a significant mediating effect between PAE and the motor and adaptive behavior domains, respectively (Table 3).

**Table 3:** Significant CpG sites (ACME *P* and TE *P* <0.05) resulting from HDMA and CMA of DNAm as a potential mediator of the association between PAE and neurodevelopmental delay at 6 months of age using the DACT method and after correction for multiple testing (BH FDR ≤0.05). ACME, ADE, TE, PM and corresponding 95% confidence intervals estimated by CMA shown.

| Domain | CpG (Gene) | ACME (95% CI) | ADE (95% CI) | Total Effect (95% CI) | Prop Med. (95% CI) |
| --- | --- | --- | --- | --- | --- |
| **Motor** | cg00086871 (TBC1D14) | -1.8 (-4.4, -0.07) | -8.1 (-15.4, -0.83) | -9.9 (-17.1, -2.9) | 0.18 (4.6e-03, 0.69) |
|  | cg00512279 (SLC18A2) | -2.5 (-6.1, -0.21) | -7.3 (-14.5, 0.46) | -9.8 (-16.7, -2.9) | 0.25 (0.02, 1.1) |
|  | cg02230180 (HNRNPU) | -2.6 (-6.1, -0.14) | -7.4 (-14.8, 0.49) | -10.0 (-16.8, -2.9) | 0.26 (0.02, 1.1) |
|  | cg03113572 (ZNF331) | -3.5 (-7.4, -0.59) | -5.9 (-12.7, 1.0) | -9.3 (-16.5, -2.2) | 0.37 (0.06, 1.3) |
|  | cg04743947 | -2.7 (-6.7, -0.03) | -6.7 (-14.3, 0.85) | -9.4 (-16.5, -2.6) | 0.29 (2.8e-03, 1.2) |
|  | cg04875987 (LRRC45) | -3.0 (-6.9, -0.29) | -6.3 (-13.5, 1.1) | -9.2 (-16.2, -2.2) | 0.32 (0.03, 1.2) |
|  | cg05141711 (AGMAT) | -2.8 (-6.7, -0.28) | -6.9 (-13.7, -0.11) | -9.7 (-16.5, -2.9) | 0.29 (0.03, 0.9) |
|  | cg09528884 (PTEN) | -4.0 (-8.2, -1.30) | -6.7 (-13.5, 5.3e-03) | -10.6 (-17.7, -3.8) | 0.37 (0.13, 0.97) |
|  | cg10137084 (CACNA2D3) | -3.3 (-7.9, -0.13) | -6.6 (-13.2, 0.99) | -9.8 (-16.5, -2.8) | 0.33 (5.6e-03, 1.3) |
|  | cg12653570 | -2.7 (-6.5, -0.19) | -7.1 (-13.5, -0.49) | -9.8 (-16.9, -2.9) | 0.27 (0.02, 0.78) |
|  | cg12789343 | -2.0 (-4.8, -0.04) | -8.0 (-15.1, -1.2) | -10.0 (-16.8, -3.1) | 0.20 (5.9e-03, 0.64) |
|  | cg19340420 (RCN3) | -2.9 (-6.1, -0.49) | -7.4 (-14.8, -0.64) | -10.3 (-17.1, -3.3) | 0.28 (0.05, 0.86) |
|  | cg22804663 (TDG) | -3.0 (-6.4, -0.09) | -6.6 (-13.9, -0.04) | -9.7 (-16.9, -2.5) | 0.31 (-3.1e-03, 0.96) |
|  | cg23238147 (TMEM51-AS1) | -2.3 (-5.4, -0.11) | -6.8 (-14.0, 0.23) | -9.2 (-16.1, -2.3) | 0.25 (3.0e-03, 1.0) |
|  | cg24716343 (PCNX2) | -2.8 (-6.2, -0.30) | -6.2 (-12.8, 1.20) | -9.0 (-16.0, -1.9) | 0.31 (0.04, 1.3) |
|  | cg26848718 (WT1) | -3.4 (-8.3, -0.23) | -5.4 (-11.9, 1.30) | -8.8 (-15.7, -2.0) | 0.39 (0.02, 1.3) |
| **Adaptive Behavior** | cg21553656 (DAG1) | -2.1 (-4.4, -0.35) | -2.3 (-5.98, 1.90) | -4.3 (-8.2, -0.3) | 0.48 (0.03, 2.0) |

The second HDMA method we tested, HIMA, did not identify any CpG sites as potential mediators of the association between either PTE and PAE and neurodevelopment at 6 months of age after correction for multiple testing.

None of the CpG sites identified as significant mediators for neurodevelopment at 6 months mediated the association between PTE or PAE and neurodevelopment at 24 months (Table S4-S5).

## Discussion

In this South African cohort, we found a significant negative association (total effect) between PTE and PAE and neurodevelopment of exposed children at 6 months of age as well as evidence of epigenetic mediation of this association using high-dimensional mediation analysis (DACT). We identified 39 CpGs that mediated the association between PTE and child cognitive (31 CpGs) and motor (8 CpGs) development. We also identified 16 CpG sites and 1 CpG site that mediated the association between PAE and child motor and adaptive behavior development, respectively. While the PTE MRS was highly predictive of PTE, it did not mediate the association between PTE and neurodevelopment. We did not find any evidence of epigenetic mediation of PTE and PAE and neurodevelopment in children of 24 months of age, most likely due to the absence of a total effect of PTE and PAE on neurodevelopment at 24 months in this sample.

Several studies have investigated the association between PTE and PAE and neurodevelopment and related disorders^51–53^. One study (N=446) conducted in a rural region of China in children with an average age of 15 months found that prenatal exposure to tobacco smoke was associated with neurodevelopmental delay in the cognitive and language domains, assessed using the BSID-III^54^. While we did find evidence of an association between PTE and cognitive development, associations with language development were not statistically significant, although it may be difficult to detect language differences as early as 6 months^41,55^. Prenatal alcohol exposure has been previously associated with decreased gross motor function, but findings have been inconsistent^56,57^. For example, a study (N=1324) that utilized the BSID-III to evaluate the relationship between PAE and gross motor development in infants of 12 months did not find evidence of an association^58^. However, the prevalence of moderate to severe PAE was lower in this cohort than in the DCHS. In our study, we found associations between PTE and PAE and neurodevelopment at age 6 months but not at age 24 months. This is in line with findings from a previous study conducted in a larger subset of the DCHS (N=734), in which PAE was associated with delayed motor development at 6 months, but not at 24 months^59^. Previous literature on neurodevelopment at 6 months of age is sparse, particularly in association with prenatal exposures. Most studies had an average infant age of at least 12 months. Unlike our present analysis, previous studies have found associations between PTE and PAE and neurodevelopment at or around 24 months of age or older^52,53,60^.

For example, a study conducted in a Polish birth cohort (N=461) identified a statistically significant association between prenatal tobacco smoke exposure and delayed development in the cognitive, language, and motor domains at 24 months of age using the BSID-III^61^. A potential explanation for this discrepancy is our relatively small sample size (N=113 at age 6 months and N=184 at age 24 months). Several of the CpG sites and associated genes that we identified through HDMA are well known for their association with sustained maternal smoking during pregnancy across two major studies from the PACE consortium^62^. Differential methylation at one of the CpG sites (cg23219570) and four genes (*MAD1L1, CAMTA1, LARP4B, FGF23*) that were identified as mediators for associations with cognitive development was previously identified in one PACE study which investigated the association of sustained prenatal smoking on changes in cord blood DNAm (450K) across 9 cohorts (N=5648)^16^. Differential DNAm at 2 CpG sites (cg02668773, cg12031962) and 7 genes (*MAD1L1, MPO, CAMTA1, COL11A2, LARP4B, ALDH1A2, SLC39A11*) in association with cognitive development and 1 CpG site (cg24671734) and 4 genes (*SKAP2, ZNF710, SLC39A11, BTBD11*) in association with motor development which we identified as mediators were previously identified in a more recent PACE study, which investigated the effects of sustained prenatal smoking on cord blood DNAm (450K) across 13 cohorts (N=6685) ^14^. It is notable that the genes *MAD1L1, CAMTA1*, and *LARP4B* had significantly, differentially methylated CpG sites across both PACE studies and our study.

Four of these genes have further been shown to be associated with cognitive outcomes. Aldehyde Dehydrogenase 1A2 (*ALDH1A2*) is associated with autism spectrum disorder (ASD) in children^63^. The *ALDH* family of genes plays a key role in the metabolism of vitamin A, an essential molecule for neuronal differentiation and development^64^. However, there has been inconsistent evidence of an association between maternal smoking and ASD^65–67^. Furthermore, brain tissue-based DNAm in two of these genes (*MAD1L1* and *COL11A1)* was associated with ASD, indicating their role in neurodevelopment ^68^.

Mitotic arrest deficient 1 like 1 (*MAD1L1)* is a key regulatory gene of the cell cycle. It has been implicated in the etiology of neurodevelopmental disorders such as SZ and BPD in several genome-wide association studies (GWAS) and candidate gene studies, including one study which found altered levels of expression of *MAD1L1* in human neural progenitor cells^69–71^. The biological mechanisms of both SZ and BPD are linked to changes in the mesolimbic reward pathway and high levels of MAD1L1 expression have been found in brain regions such as the ventral tegmental area^72^. Previous studies have found that maternal smoking was associated with a higher rate of SZ and BPD in offspring, suggesting that differential expression of MAD1L1 may be a plausible pathway that links smoking to neurodevelopmental disorders^73^.

Calmodulin-binding transcription activator 1 (*CAMTA1*) is also involved in cell-cycle regulation and is highly expressed in the brain^74^. Studies have found that *CAMTA1* plays a role in memory production and recall, dysfunction of which is implicated in disorders such as down syndrome, schizophrenia, and depression^75,76^. However, these studies have been conducted in older adults, and more research needs to be done to corroborate memory dysfunction in children with differential expression of *CAMTA1*.

Two genes of note that we identified as significant mediators of the association between PAE and motor development are *SLC18A2* and *HNRNPU* ^77^. Homozygous mutations in *SLC18A2*, a vesicular monoamine transporter, have been implicated in the presentation of infantile movement disorder also known as infantile hypotonic parkinsonian disorder or brain dopamine-serotonin vesicular transport disease^78–80^. Mutations in *HNRNPU* have been associated with infantile spasms^81,82^. However, there has been little to no research published on prenatal and early life exposures or pediatric movement disorders and related epigenetic modifications.

Methylation risk scores can be used as a potential biomarker for prenatal smoking and alcohol use and to reduce the dimensionality of methylation data, giving us more statistical power to detect associations^42^. Although our methylation risk score for prenatal smoking was highly predictive of PTE, it did not significantly mediate the association between PTE and neurodevelopment. One possible explanation is that our MRS algorithm only considers the relationship between exposure and mediator, whereas HDMA considers both the E-M and M-O relationships. Additionally, it is possible that the difference in ancestry between the external studies (primarily European ancestry) and the DCHS (African and mixed ancestry) reduced the prediction accuracy of our MRS for both prenatal smoking and prenatal alcohol use^83^.

Our study has a few limitations. First, unlike PTE which has been extensively studied^14–16,84^ in association with DNAm, the association between PAE and DNAm is not well understood, which makes the comparison of our findings to existing literature more challenging. The vast majority of epigenetic studies evaluating the effects of prenatal alcohol exposure are conducted in birth cohorts from HICs and tend to have a low prevalence of PAE^17,62^. Because of the low prevalence of the exposure, these studies often do not have the statistical power to detect an association. A PACE study that investigated the association between PAE and cord blood DNAm (450K) across 6 cohorts (N=1147) did not find strong evidence of a relationship^17^. This limitation also in part explains the poor performance of our methylation risk score for PAE. Furthermore, although the DCHS has a higher prevalence of both PAE and PTE compared to HIC cohorts, this study had a relatively small sample size at both 6 months (N=113) and 24 months (N=184), which may have limited our statistical power to detect associations between PTE and PAE and neurodevelopment across other domains and at 24 months of age.

Another limitation to our study is the possible presence of exposure misclassification bias. Underreporting is a common problem when attempting to ascertain socially undesirable behaviors such as smoking or alcohol consumption during pregnancy. In our case, alcohol use was measured using a self-report questionnaire and smoking was measured via urine cotinine levels. Although using cotinine as a biomarker of smoking is not affected by social desirability bias, it was only measured at one time point in this cohort. Exposure misclassification can be an issue in epigenomic mediation analyses in that the DNA methylation estimates may be a more accurate indicator of exposure than the variables available in our cohort, which can lead to an overestimation of the mediation effect^85^.

Furthermore, while blood is the most common tissue used to evaluate the epigenomic mechanisms of a multitude of risk factors and outcomes, it is not yet clear whether blood tissue can be effectively used to evaluate neurological outcomes. Epigenetic signatures are tissue and cell-type specific, and as such, the selection of relevant tissues and cell-type is crucial for epigenetic studies. While brain tissue would be the most relevant tissue to study in association with neurodevelopment, cord blood has the advantage in that it is more easily accessible from living individuals and could therefore be linked to future cognitive outcomes^86^. Additionally, epigenetic signatures in cord blood may represent systematic and mechanistic changes through various peripheral pathways or may show blood-brain concordance, and thus are important biomarkers of neuropsychiatric disease^87^.

Our study has several strengths. To start, we have data at 6 months of age, which may allow us to infer a causal relationship more confidently because of the short temporality between birth and measurement. Further, while this analysis had a small sample size, the use of novel methods to increase our statistical power allowed us to detect more potentially meaningful associations and is an important strength. To our knowledge, this is one of the first epigenomic studies to employ HDMA. The HIMA method, which utilizes the joint significance test, was potentially too conservative and underpowered to detect significant mediators in this study^45^. DACT has been shown to be more powered than HIMA. The use of methylation risk scores and high-dimensional mediation analysis in the field is still in its infancy. We recognize that more studies will be required to validate the robustness of these methods as the field advances, but this study provides some insight into the utility and potential of HDMA in real-life settings.

Finally, our analysis was conducted on a subset of a well-characterized, underrepresented, birth cohort from an LMIC. We were able to detect associations due to the high prevalence of behavioral risk factors -such as maternal smoking and alcohol consumption-in such vulnerable populations, a characteristic that cohorts from HICs typically do not exhibit. This highlights the importance of conducting large-scale multi-ethnic cohort studies to advance the field of (epi)genomics and reduce health disparities. As Wojcik et al. point out, while the burden of disease disproportionality lies with marginalized populations, the majority of studies are conducted in populations of European ancestry, which limits the field’s ability to investigate the burden of disease in vulnerable communities^88^. As follows, investigating epigenomic associations with health outcomes in a well-characterized cohort from an LMIC allows us to add to the sparse literature and shed light on the importance of prioritizing cohorts like the DCHS.

Overall, these findings suggest that epigenetic changes in the methylome may in part explain the biological mechanisms underlying the relationship between maternal smoking and alcohol consumption during pregnancy and child neurodevelopment across the cognitive, motor, and adaptive behavior domains. Our study provides motivation to conduct larger mediation studies to replicate our findings. Mediation analyses in epigenomics are important for discerning causal mechanisms from exposure to disease. The identification of significant CpG sites could provide novel insights for the early detection of disease and potential prevention targets in translational research and community interventions in at-risk populations.

## Data Availability

All data produced in the present work are available upon reasonable request to the authors.

## Acknowledgements

The authors would like to thank the study and clinical staff at Paarl Hospital, Mbekweni and TC Newman clinics, as well as the CEO of Paarl Hospital, and the Western Cape Health Department for their support of the study. The authors also thank the families and children who participated in this study.

