## Supplementary Material for "DNA Methylation as a Potential Mediator of the Association Between Prenatal Tobacco and Alcohol Exposure and Child Neurodevelopment in a South African Birth Cohort"

### Supplementary Methods

#### **Socioeconomic status (SES) composite variable**

The SES composite variable was calculated as follows:

**INPUTS**

STEP 1: RESPONSE CODING

a. Maternal employment
   i. Employment = 1 vs Unemployment = 0
b. Maternal education
   i. Primary education = 0
   ii. Some secondary = 1
   iii. Completed secondary = 2
   iv. Any tertiary = 3
c. Household income
   i. <R1000/m = 0
   ii. R1000 – 5000/m = 1
   iii. >R5000/m =2
d. Household assets and financial activities
   i. Sum over 10 items (each checked item counts as 1):
      1. Electricity
      2. Tap or running water
      3. Domestic worker
      4. A flush toilet inside
      5. A built-in kitchen sink
      6. An electric stove or hotplate
      7. A working telephone (this includes a cell phone)
      8. At least one motor car or truck
      9. A motorcycle or scooter
      10. A bicycle
   ii. Sum over 3 items (each checked item counts as 1)
       1. Shop at supermarkets
       2. Use any financial services (such as bank account, ATM card or credit card)
       3. Have an account at a retail store (e.g. Pep, Jet, etc)

STEP 2: STANDARDIZATION OF INPUTS

a. Standardize the sum of household asset/financial activities (variable name: ***“stdassetsum”***)
b. Standardize income (variable name: ***“stdincome”***)
c. Standardize education (variable name: ***“stdeducation”***)

**OUTPUTS**

STEP 1: DERIVING A *CONTINUOUS* SOCIOECONOMIC COMPOSITE

a. Continuous socioeconomic status composite (variable name: *“****sessumscore****”*)

**Sessumscore** = **stdassetsum** + **stdeducation** + **stdincome** + **(employment*0.5)**

STEP 2: DERIVING A *CATEGORICAL* SOCIOECONOMIC COMPOSITE

b. Categorical socioeconomic status composite (variable name: *“****sesquartile****”*)

**Sesquartile** is derived from **sessumscore** composite split into **quartiles**:

1. Lowest SES
2. Low-mod SES
3. Mod-high SES
4. High SES

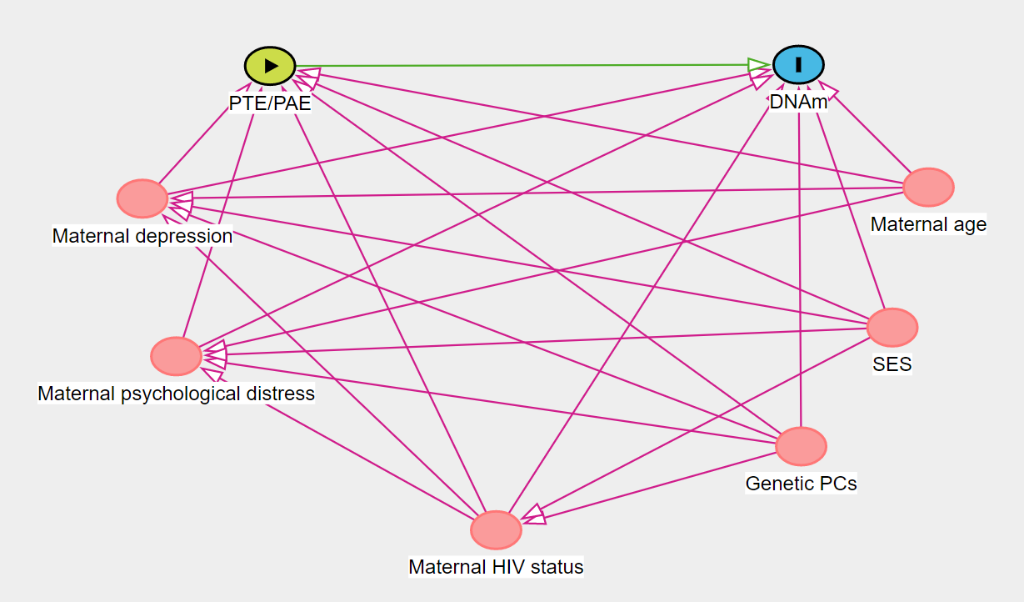

**A**

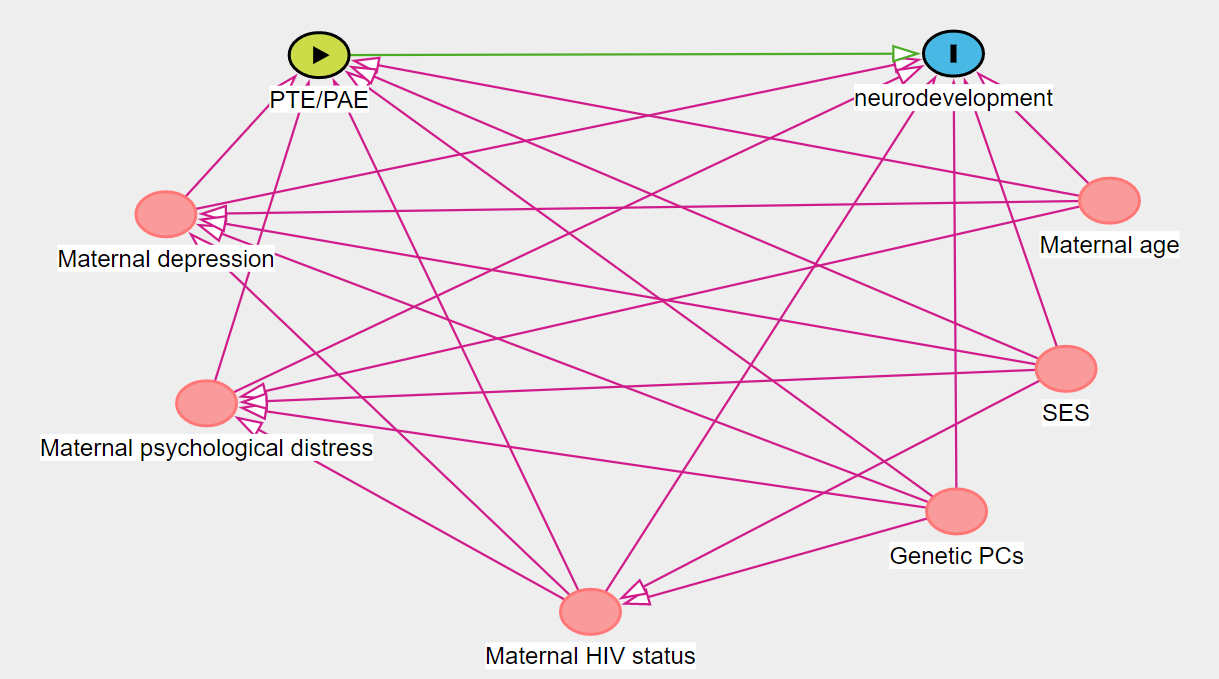

**B**

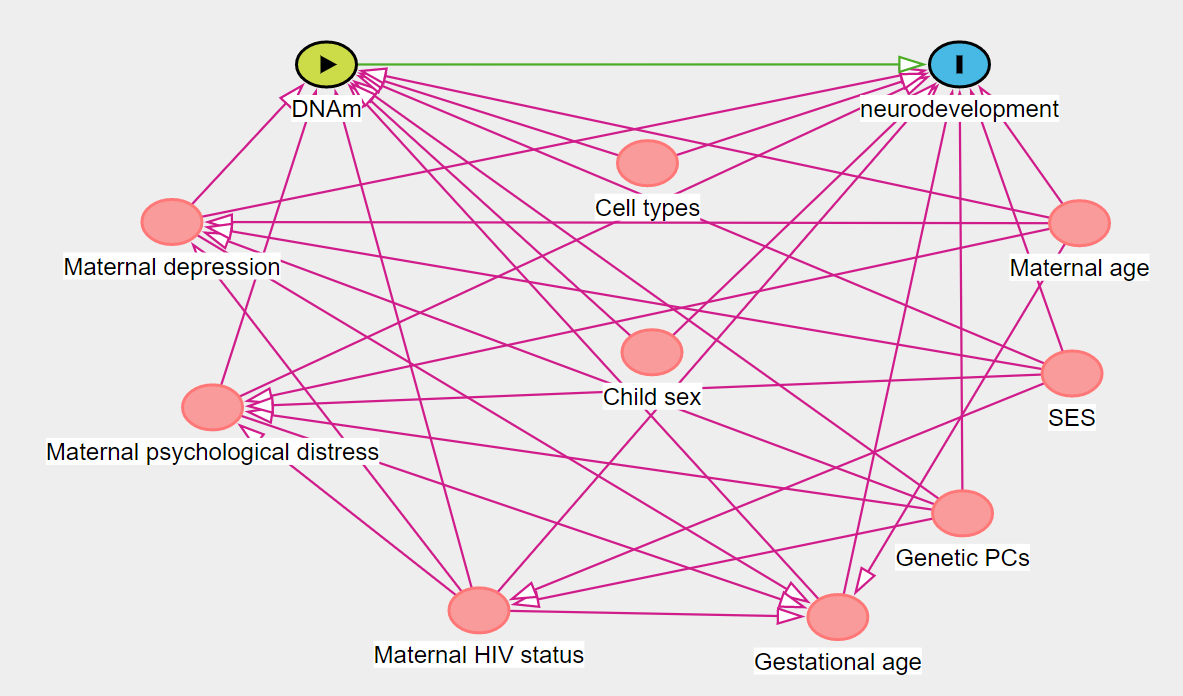

**C**

**Figure S1:** Directed acyclic graphs (DAGs) used for confounding assessment. **S1A)** Exposure-mediator DAG. **S1B)** Exposure-outcome DAG. **S1C)** Mediator-outcome DAG

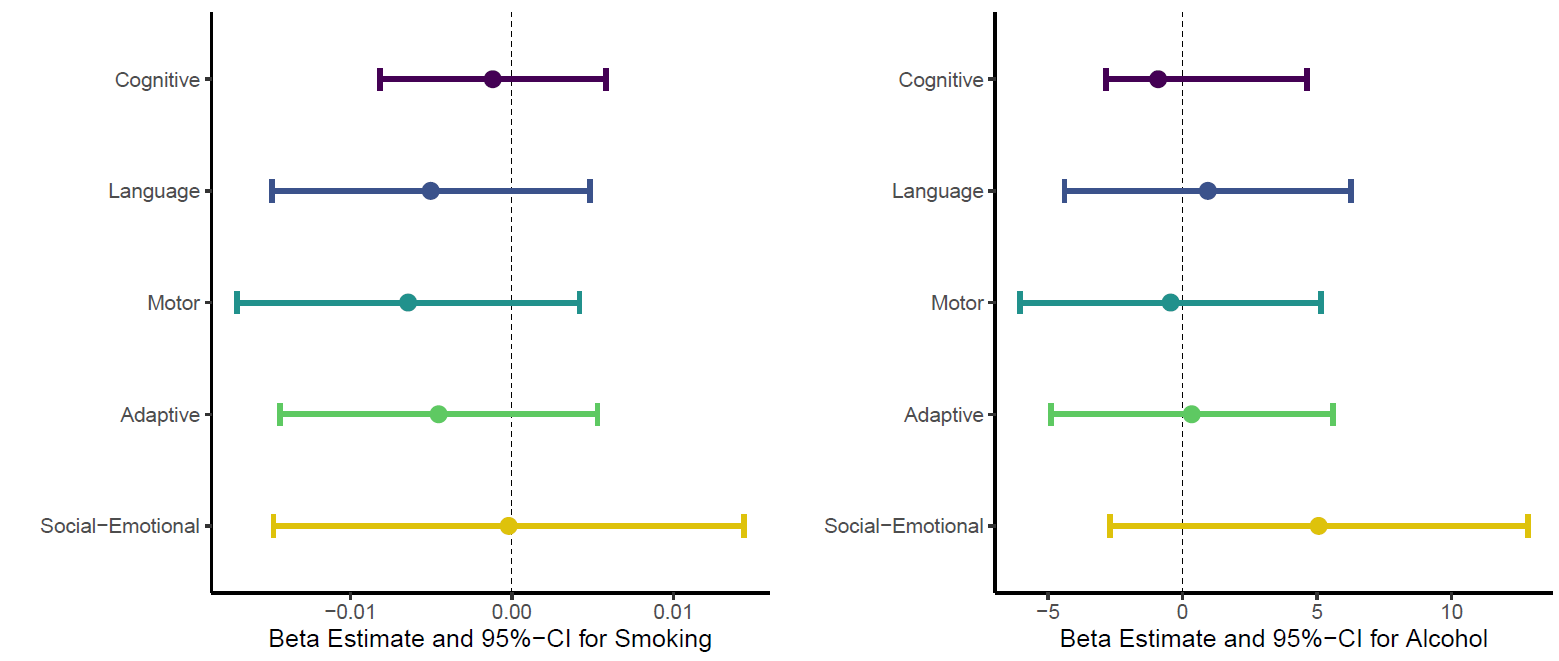

**B**

**A**

**Figure S2:** Association of PTE and PAE on neurodevelopment domains at 24 months. Models were adjusted for paternal SES, maternal depression, maternal psychological distress, gestational age, maternal age, maternal HIV status, cell-type proportions, and the first 5 genetic principal components. **S2A)** Association of PTE on neurodevelopment domains, additionally adjusted for PAE. **S2B)** Association of PAE on neurodevelopment domains, additionally adjusted for PTE.
